# The global burden of pressure ulcers among patients with spinal cord injury: a systematic review and meta-analysis

**DOI:** 10.1101/19007237

**Authors:** Wondimeneh Shibabaw Shiferaw, Tadesse Yirga, Henok Mulugeta, Yared Asmare Aynalem

**Affiliations:** Lecturer of Nursing, Department of Nursing, Institute of Medicine and College of Health Science, Debre Berhan University; Lecturer of Nursing, Department of Nursing, College of Health Science, Debre Markos University

**Keywords:** Pressure ulcers, spinal cord injury, Systematic review, Meta-analysis, Ethiopia

## Abstract

**Background:** Pressure ulcer, one of the common challenging public health problems affecting patient with spinal cord injury, is the formation of lesion and ulceration on the skin specially in the bony prominence areas. It has a significant impact to the patient and health care system. Moreover, it has psychological, physical, social burden and decrease the quality of life (QoL) of patients. Despite its serious complications, limited evidence is available on the global magnitude of pressure ulcers among patient with spinal cord injury. Hence, the objective of this systematic review and meta-analysis was to estimate the global magnitude of pressure ulcers among patient with spinal cord injury.

**Methods:** PubMed, Scopus, Google Scholar, Africa journal online, PsycINFO and web-science were systematically searched online to retrieve related articles. The Preferred Reporting Items for Systematic Review and Meta-Analysis (PRISMA) guideline was followed. The random-effects model was fitted to estimate the summary effect. To investigate heterogeneity across the included studies, I^2^ test was employed. Publication bias was examined using funnel plot and Egger’s regression test statistic. All statistical analysis was done using STATA version 14 software for windows.

**Results:** Twenty-four studies which comprises of 600,078 participants were included in this meta-analysis. The global pooled magnitude of pressure ulcer among patients with spinal cord injury was 32.36% (95% CI (28.21, 36.51%)). Based on the subgroup analysis, the highest magnitude of pressure ulcer was observed in Africa 41.19% (95% CI: 31.70, 52.18).

**Conclusion:** This systematic review and meta-analysis revealed that about one in three patients with spinal cord injury had pressure ulcers. This implies that the overall global magnitude of pressure ulcer is relatively high. Therefore, policymakers (FMoH) and other concerned bodies need give special attention to reduce the magnitude of pressure ulcers in patient with spinal cord injury.

## Background

Spinal cord injury (SCI) is a life-threatening and debilitating injury with tremendous immediate and long-term extensive impact on the medical, social, psychological and economic aspects of clients, their caregivers and the society [1-3]. The annual incidence rate of SCI is 44 cases per 1 000 000 people in Tehran [4], and in European countries ranges from 5.5 to 195.4 cases per million inhabitants [5]. Spinal cord injured patients have a high risk of developing pressure ulcers due to motor and sensory impairments, immobility, changes in skin composition, and prolonged hospital stay [6, 7]. PrUs are a serious, costly, and life-long complication of SCI. Around, 30–40% of clients with spinal cord injuries develop pressure ulcers during the acute and rehabilitation phases, most commonly over bony prominences[8].

Pressure ulcers and their treatment represent one of the most challenging clinical problems faced by patients who are neurologically impaired or have chronic spinal cord injury (SCI) [9]. Even though, there are different pressure ulcer classification systems, multiple sources of data, and varying methods of obtaining data. According to the National Pressure Ulcer Advisory Panel (NPUAP) consensus development conference, pressure ulcers are classified according to severity from suspected deep tissue injury through Unstageable, with suspected deep tissue injury representing the earliest stage of pressure ulcer formation, and Unstageable is defined as “full thickness tissue loss in which the base of the ulcer is covered by slough (yellow, tan, gray, green, or brown) and/or eschar (tan, brown, or black) in the wound bed”[10].

Pressure ulcers have a significant impact to the affected individual and on the health care system. It highly affects the psychological, physical, social well-being and the quality of life of the affected individuals [11-13]. Likewise, it lead to recurrent hospitalizations, multiple surgeries, potentially devastating complications, morbidity and early mortality [9, 14, 15]. For example, a study done in Canada revealed that the economic burden of pressure ulcer among spinal cord injury was an average of $18,758 [3]. Pressure ulcer may account for 25% of the overall cost of treating paraplegic and tetraplegic persons [16]. Moreover, a study done in Canada showed average monthly cost per community dwelling SCI individual with a PU was $4745 [17].

The development of pressure ulcers (PUs) during hospitalization of patients with a spinal cord injury (SCI) has been reported in different literature, which varying from 11% to 50% in the current publications[6, 18]. Similarly, a study done in Switzerland reported that the incidence of hospital acquired pressure ulcer was 2.31 per patient-year [18]. Poor pressure relief practices lead to PU development in persons with SCI [10]. Management and care of pressure ulcer has become a serious public health challenge, with longer hospital stays than for other causes. Preventive measures to decrease the development of pressure ulcers consisted of basic skin care, pressure dispersion using fenestrated foams and alternating weight-bearing sites by regular frequent positioning and turning[7]. In addition, the key targets for interventions has been advocated to reduce the burden of pressure ulcers in patient with SCI. These interventions includes identification of risk factors, patient education, acute intensive care, and support body surfaces [19].

The development of PUs is a very complex phenomenon and due to the presence of multiple risk factors. Identifying risk factors used as benchmarks to design appropriate prevention measure, to improve client safety and efficient utilization of resources. Several risk factors are responsible for the development of pressure ulcers in patient with spinal cord injury. For instance, duration after SCI(> 1 year) [20], age(older age), sex(being male) [20, 21], poor nutritional status [22],quadriplegia[23-25],smoking[6, 26],comorbidity[23, 27], severe Braden scores[28], weight(being underweight)[26], lower level of education[20, 21], and lack of an intimate partner[21] were some of the risk factors associated with PU. Similarly, it has been reported that patients with higher-level spinal cord injuries are more susceptible than those with lower-level lesions [13].

Pressure ulcers among spinal cord injured client’s remains unrelenting problem and is a major issue in nursing care across the globe. Prevention of pressure ulcers is one of the key components of nursing care, and one of the quality indicator of nursing care at large [29]. Although most previous studies have been conducted to assess the magnitude of pressure ulcer in acute care setting, in intensive care unit, and on public hospitals, the global magnitude of pressure ulcers among patient with spinal cord injury remains unknown. Hence, this study aimed to estimate the global burden of pressure ulcers among spinal cord injured patients. Finding from the current study would serve as benchmark for policy-makers to implement appropriate preventive measure and to alleviate the pressing problem of pressure ulcer. In addition, for clinicians estimating burden of pressure ulcer would reflect overall quality indicator for facilities and a way to assess the efficiency of prevention strategies.

## Methods

### Search strategy and database

A two-step search strategy was used to identify all relevant literature. First, six electronic databases such as PubMed, Google Scholar, Africa journal of online, Scopus, Web-science, and PsycInfo were searched to extract all available literature. Second, a hand search of gray literature and other related articles in order to identify additional relevant research. In addition, all electronic sources of information were searched from 1^st^ January/ 2000 to 1^st^ July2, / 2019. The search was conducted using the following MeSH and free-text terms: “pressure ulcer”, “pressure injury”, “decubitus ulcer”, “spinal cord injury”, and “prevalence”. Boolean operators like “AND” and “OR” were used to combine search terms.

### Eligibility criteria

Studies were included if they met the following criteria: (1) All observational studies, which reported the prevalence pressure ulcer among spinal cord injured clients;(2) articles published in peer reviewed journals and gray literature:(3) published in the English language between 2000 to 2019; (4) we imposed no restriction on the area where it is conducted; and (5) the group of patients admitted without PUs. Studies were excluded on any one of the following conditions:(1) Patients admitted with pressure ulcer;(2) studies with poor quality score as per stated criteria; and (3) articles in which fail to determine the outcome (pressure ulcer).

### Selection and quality assessment

Data were extracted by three authors using a pre-piloted and standardized data extraction format prepared in a Microsoft excel. Data extracted from articles included authors name, year of publication, study area, study design, sample size, prevalence and data collection methods. The quality of each included study was assessed using Newcastle-Ottawa scale [30]. Studies were included in the analysis if they scored ≥7 out of 9 points in three domains of the equally weighted modified NOS components for cohort studies and for cross-sectional studies quality assessment tool, score of ≥5 out of 10 considered as high quality score. Finally, the quality score of each study were extracted from each incorporated article by three independent authors. Any disagreements at the time of data abstraction were resolved by discussion and consensus. Additional file 1: Table S1. Methodological quality assessment of cross-sectional studies using modified Newcastle - Ottawa Scale (NOS). Table S2. Methodological quality assessment of cohort studies using modified Newcastle - Ottawa Scale (NOS).

### Statistical analysis

To obtain the pooled effect size, a meta-analysis using weighted inverse variance random-effects model was performed. Heterogeneity across the included studies was checked using the I^2^ statistics test[31]. In addition, to investigate the possible sources of heterogeneity, meta-regression analysis was deployed. Publication bias was assessed by visual inspection of a funnel plot. Similarly, Egger was conducted and a p<u>□□≤ 0</u>.05was considered statistically significant for the presence of publication bias [32, 33]. Moreover, sensitivity analysis was performed to investigate whether the pooled effect size was influenced by individual studies. The data were analysed using STATA version 14 statistical software[34].

### Data synthesis and reporting

We analysed the data to estimate the pooled magnitude of pressure ulcers among spinal cord injured clients. Results were presented using forest plots. Preferred Reporting Items for Systematic Reviews and Meta-Analyses (PRISMA) guideline was followed to report our results[35]. (Supplementary file 2-PRISMA checklist) and, it is not registered in the Prospero database.

## Result

### Search results

We found that a total of 1,053 articles, of these, 1,027 studies were found from six international databases and the remaining 26 were through manual search. Databases includes; PubMed (611), Scopus (171), PsycInfo (10), Google scholar (76), Web-science (92), and Africa online journal (67). Out of them, 529 duplicate records were identified and removed. From the remaining 524 articles, 407 articles were excluded after reading of titles and abstracts based on the pre-defined eligible criteria. Finally, 117 full text articles were assessed for eligibility criteria. Based on the pre-defined criteria and quality assessment, only 24 articles were included for the final analysis (Figure 1).

**Figure 1.**
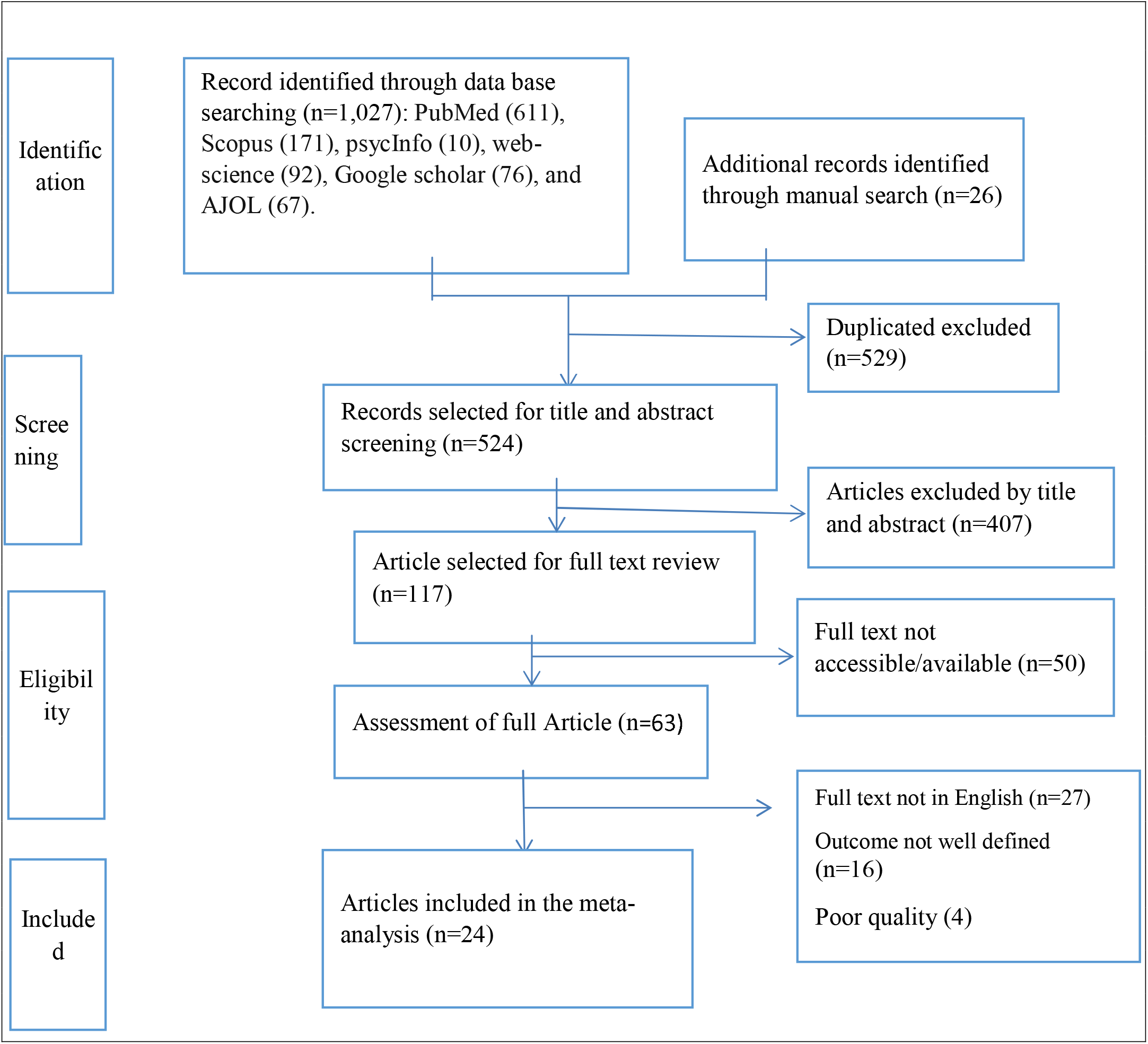
PRISMA flowchart diagram of the study selection.

## Baseline characteristic of the included studies

A total of 24 studies with 600,078 participants were included in this meta-analysis. Overall information regarding the prevalence was obtained from various areas across a globe: 8 studies from America [6, 36-41], 7 article from Europe [18, 23, 42-46], 5 research from Asia[20, 28, 47-49] and 4 studies from Africa [7, 22, 24, 50]. The highest prevalence of pressure ulcers (56%) was reported from Europe and the lowest (11%) in America. Concerning sample size, the number of study participants ranges from 38 to 7489. Moreover, based on modified Newcastle Ottawa quality score assessment all 24 articles fulfil the required quality score (Table 1).

**Table 1.** demonstrates the baseline characteristic of primary studies.

| **First Author** | **Pub. year** | **study area, continent** | **study design** | **sample size** | **Prevalence% (95%CI)** | **Data Collection year** | **data collection methods** | **Quality score** |
| --- | --- | --- | --- | --- | --- | --- | --- | --- |
| Ash, D etal [42] | 2002 | United Kingdom, Europe | Cross-sectional | 144 | 56(47.9,64.1) | 1998 to 2000 | document review | 7 |
| Brienza,D., et al[36] | 2017 | United States,North America | cohort | 104 | 37.5(28.2,46.8) | 2008–2012 | observation and examination | 7 |
| Chopra,T.,et al[51] | 2016 | United States, North America | Cross-sectional | 201 | 38(31.3,44.7) | January 2004 and December 2008 | document review | 7 |
| DeJong, G., et al.[37] | 2014 | United states, north America | Cohort | 159 | 13.1(7.8,18.3) | NA | Document review | 7 |
| Eslami V et al[20] | 2012 | Iran , Asia | Cross-sectional | 7,489 | 34.6(33.5,35.7) | June 2007 to June 2009 | physical examination | 9 |
| Fazel FS, etal [52] | 2018 | Iran,Asia | cohort | 580 | 28.1(24.4,31.7) | June 2013 to December 2015 | Observation and examination | 8 |
| Garber, S.L., et al [38] | 2000 | United states, North America | Cohort | 118 | 31(22.2,39.3) | NA | Interview and exam | 8 |
| Haisma, J.A., et al [43] | 2007 | Netherlands, Europe | cohort | 212 | 36(29.5,42.5) | May 2000 and September 2003 | Self-report | 8 |
| Idowu, O., et al [22] | 2011 | Nigeria,Africa | cohort | 105 | 45.9(36.4,55.4) | 1 October 2004 and 30November 2006 | Skin examination | 8 |
| Iyun A.O. etal [7] | 2012 | Nigeria, Africa | cohort | 67 | 47.7(35.7,59.7) | January 2003 to June 2004 | Self-report and documentation | 7 |
| Joseph, C. and L.N. Wikmar [24] | 2015 | south Africa,Africa | cohort | 141 | 29.8(22.2,37.4) | 15 Septem 2013 to 14 Septem 2014 | observation and examination | 8 |
| Kovindha, A. et al[47] | 2015 | Thailand, Asia | Cross-sectional | 129 | 26.4(18.8,34) | 1 January 2013 to 31December 2013 | Self-report | 7 |
| Krishnan,S., et al [39] | 2017 | United States, North America | Cross-sectional | 3,098 | 20.3(18.9,21.7) | 1993 to 2006 | document review | 8 |
| Li ,C., et al[6] | 2016 | United States,North America | Cross-sectional | 350 | 11(7.6,14.4) | August 2011 to February 2014 | Self-report | 8 |
| Löfvenmark I et al[50] | 2016 | Botswana,Africa | cohort | 38 | 48(32.1,63.9) | 1February 2011 to 31January 2013 | Skin examination | 8 |
| Raghavan, P., et al.,[44] | 2003 | United kingdom,Europe | Cross-sectional | 472 | 23(19.2,26.8) | NA | Observation and exam | 7 |
| Richard-Denis, A., et al [40] | 2016 | Canada,North America | Cohort | 123 | 33.3(24.9,41.6) | January 1, 2009, and December 31, 2011 | Document review | 7 |
| Saunders et al[41] | 2013 | United States, North America | Cross-sectional | 2549 | 19.9(18,21.7) | NA | mail-in survey | 9 |
| Scheel-Sailer, A., et al [45] | 2013 | Switzerland, Europe | cohort | 185 | 25.4(19.1,31.7) | 1 Septem 2009 to 28February 2010 | observation and examination | 7 |
| Sheerin, F. et al[46] | 2005 | Ireland, Europe | Cross-sectional | 82 | 37(26.5,47.4) | December 2000 to December 2002 | document review | 7 |
| Taghipoor, K.D., et al [49] | 2009 | Iran, Asia | Cross-sectional | 3791 | 39.1(37.5,40.6) | NA | Document review | 7 |
| Tchvaloon,E., et al[48] | 2007 | Israel, Asia | cohort | 143 | 26.6(19.4,33.8) | 1962 and 2004 | document review | 7 |
| van der Wielen H et al[18] | 2016 | Switzerland, Europe | cohort | 185 | 50(42.8,57.2) | September 2009 to February 2010 | observation | 7 |
| Verschueren J et al[23] | 2011 | Netherlands, Europe | cohort | 193 | 36.5(29.7,43.3) | NA | observation and examination | 8 |

N/A :not applicable

### Global burden of pressure ulcer among patient with spinal cord injury

The result of this meta-analysis using random effects model showed that the global pooled magnitude of pressure ulcers among spinal cord injured clients were 32.36% (95% CI: 28.21-36.51) (Figure 2) with high significant level of heterogeneity was observed (I^2^ = 97.1%; p<0.001).

**Figure 2.**
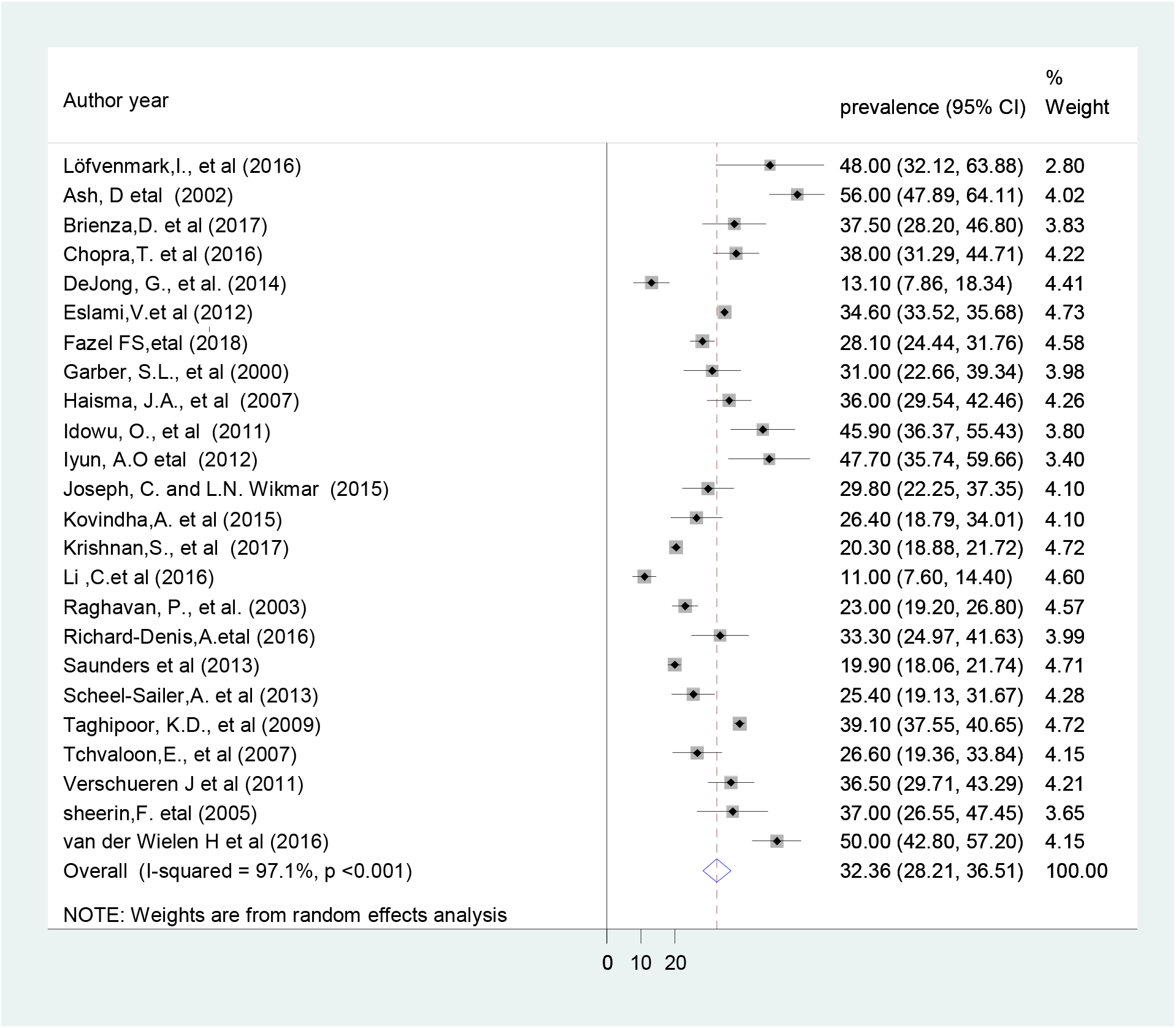
The global pooled prevalence of pressure ulcer among spinal cord injured clients.

### Sub-group analysis

In order to validate the presence of significant heterogeneity within and between the primary studies require the need to conduct subgroup analysis. As a result, the finding of subgroup analysis using study design showed that the highest magnitude of pressure ulcer was observed among studies done using cohort study design which is 34.85% (95% CI: 28.50, 41.19), I^2^=88.4%) (Figure 3). Concerning with study area, high burden of pressure ulcers was observed among studies done in Africa which is 41.94 %(95% CI: 31.70, 52.18), I^2^ =72.7%) (figure4).

**Figure 3.**
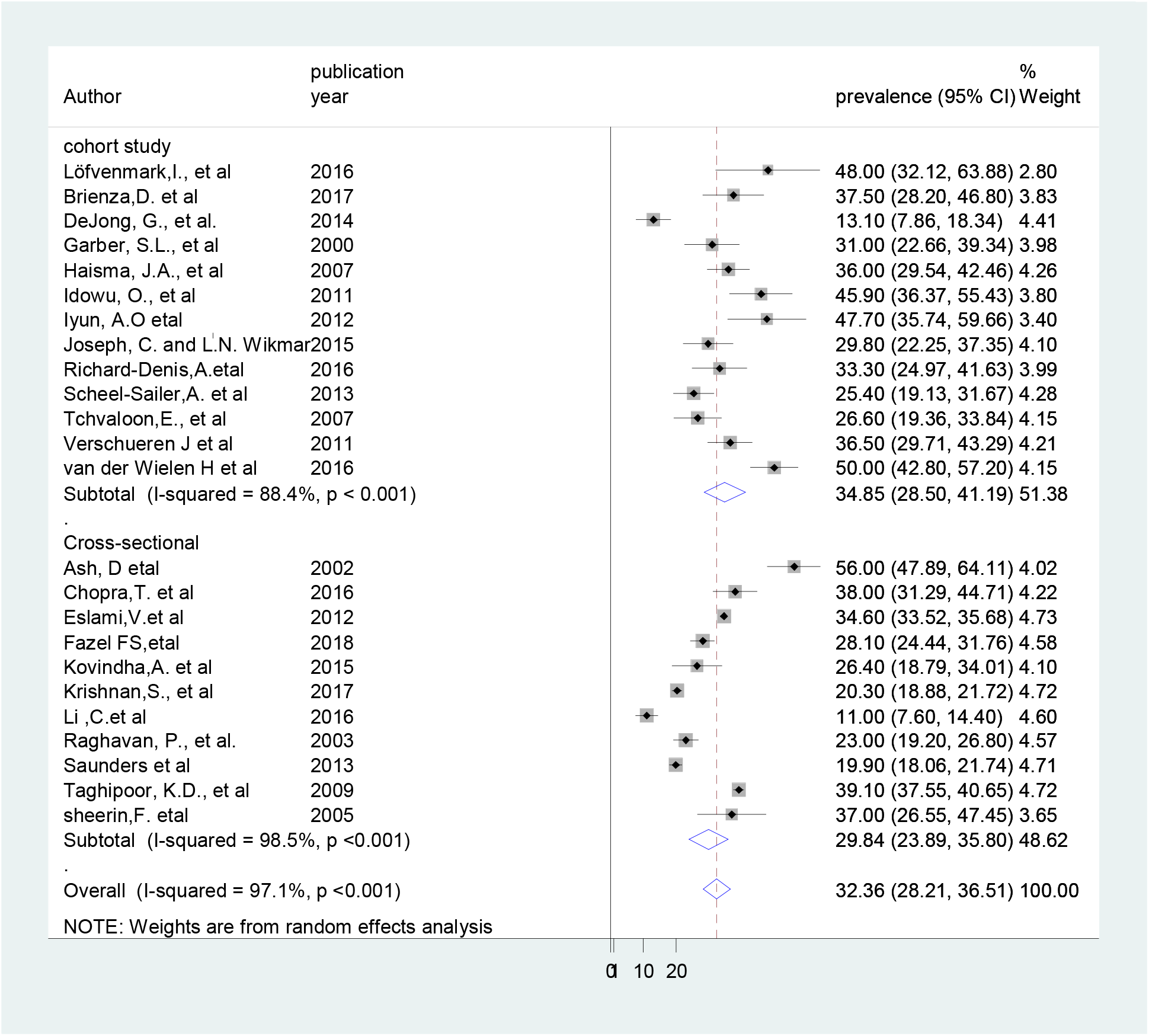
Forest plot showing subgroup analysis by study design.

### Meta-regression analysis

To investigate the possible source of variation across the included studies, we have performed meta-regression by using publication year, and sample size as covariate of interest. However, the result of the meta-regression analysis showed that both covariates were not statistically significant for the presence of heterogeneity (Table 2).

**Table 2.** Meta regression analysis for the included studies to identify source of heterogeneity

| Covariate (source) | Coef. | Std. err. | P-value | 95% Conf. Interval |
| --- | --- | --- | --- | --- |
| Publication year | -0.416 | 0.530 | 0.442 | -1.526, 0.694 |
| Sample size | 0.0002 | 0.001 | 0.869 | -0.003, 0.0035 |

### Publication bias

To identify the presence of publication bias, funnel plot, and egger’s test was performed. The visual inspection of the funnel plots showed asymmetrical distribution, which is the evidence for publication bias (Figure 5). However, asymmetry of the funnel plot was not statistically significant as evidenced by egger test(P=0.74).

**Figure 4.**
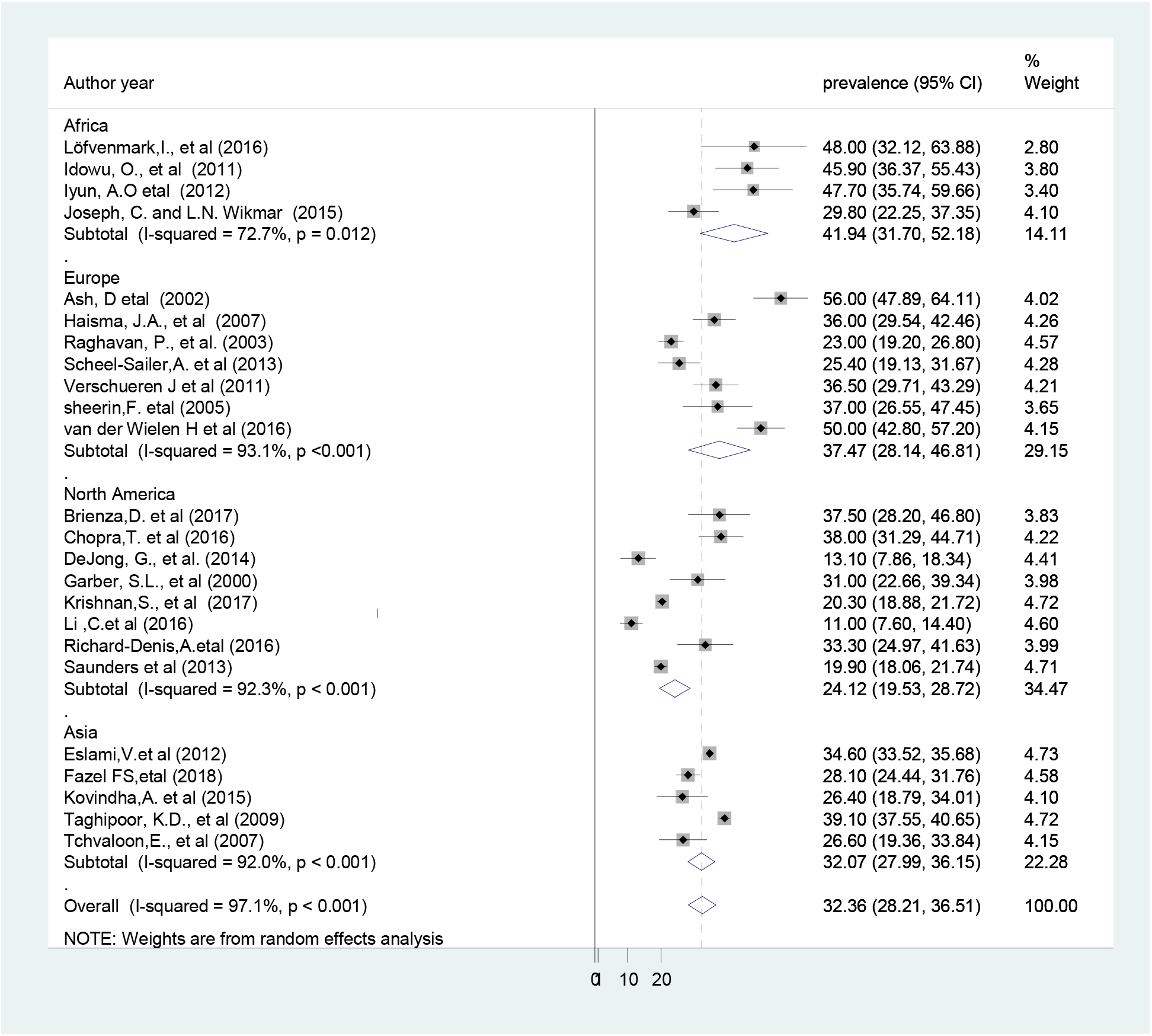
Forest plot showing subgroup analysis by study continent.

**Figure 5.**
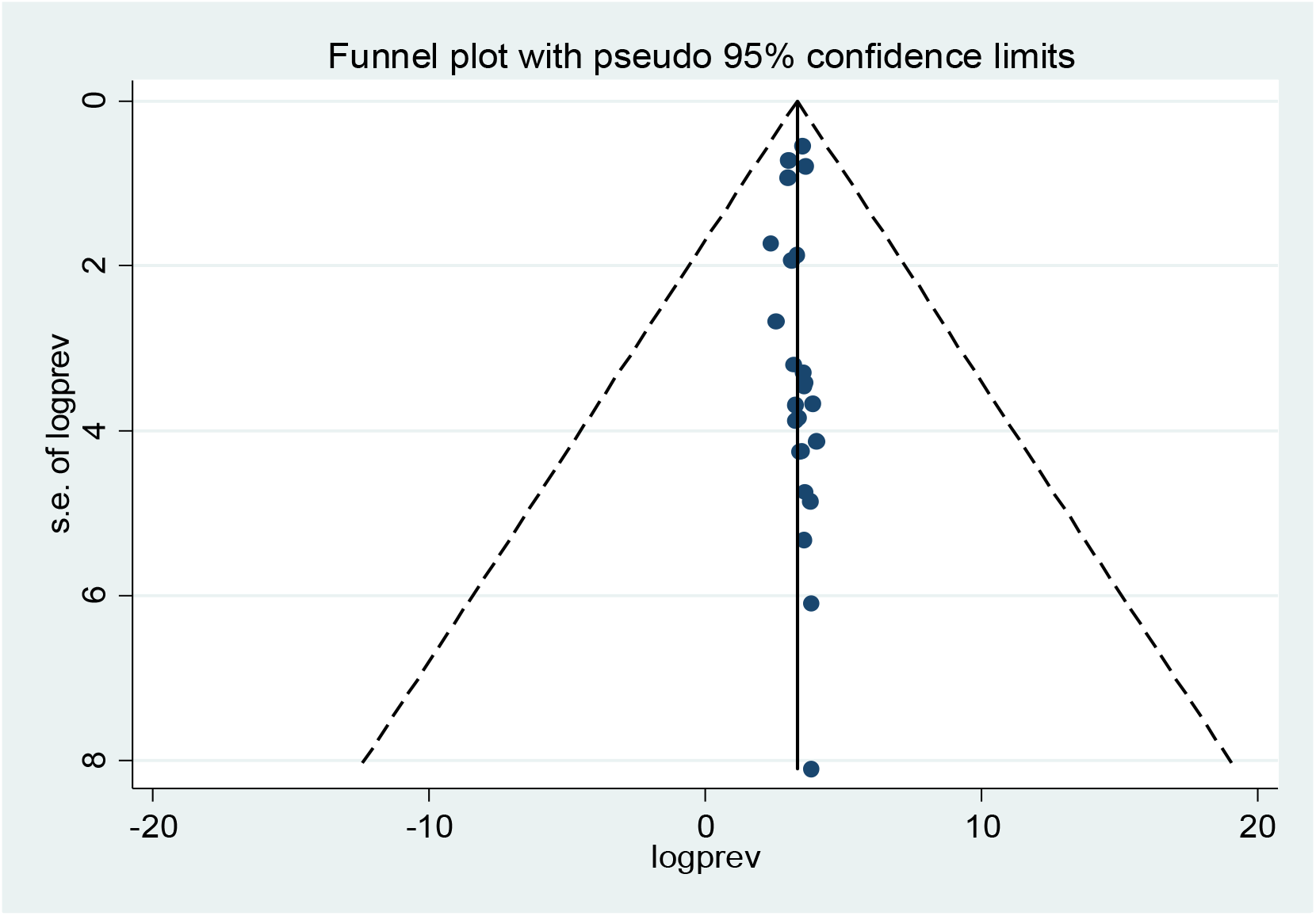
Funnel plot to test the presence of publication bias of the 24 studies.

### Sensitivity analysis

We have also conducted sensitivity analysis, to evaluate the effect of individual study on the pooled effect size. The finding of sensitivity analyses using random effects model revealed that no single study affect the overall magnitude of pressure ulcer (figure 6).

**Figure 6:**
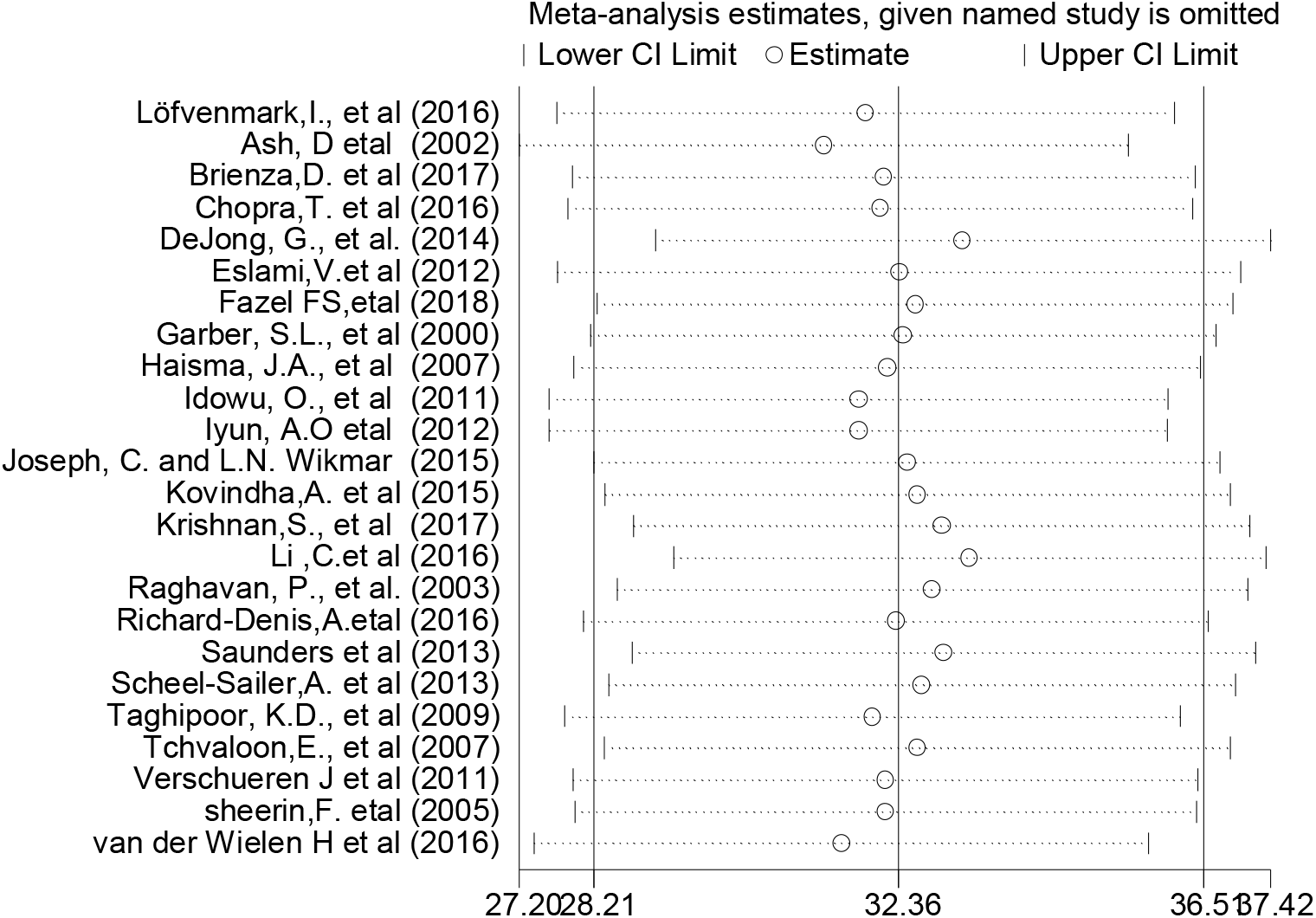
Result of sensitivity analysis of the 24 studies.

## Discussion

This study aimed to synthesis evidence on the magnitude of pressure ulcer among spinal cord injured patients. This meta-analysis provides a comprehensive estimation of the global pooled prevalence of pressure ulcer. In line with the objective, the present study found that the overall prevalence of pressure ulcer among spinal cord injured patient was 32.36% (95% CI: 28.21, 36.51%). This finding is much higher than a meta-analysis study done on the global prevalence of pressure ulcer in public hospitals 14.8% [53]. This showed that there is a high magnitude of pressure ulcer among patients with spinal cord injury, which indicates inadequate prevention and management of pressure ulcer risk factors.

The result of the subgroup analysis based on study design (cross-sectional vs cohort) showed that the highest pooled prevalence of pressure ulcers was observed from studies done with cohort design 34.85% (95%CI: 28.50, 41.19). This variation might be in case of cohort study design the outcome variable was obtained through observation, skin assessment, physical examination and with certain follow up time whereas in cross-sectional studies data were collected with document review and self-report. Therefore, these situations may contribute for variation across study designs. Similarly, we had performed subgroup analysis based on continent, the highest magnitude of pressure ulcer reported in Africa 41.94%, followed by Europe 37.47%. The possible explanations for this variation might be due methodological differences, variation in quality of care, policy and strategy difference[9, 54].

This meta-analysis has implication for clinical practice. Estimating the burden of pressure ulcers provide current evidence to establishing robust preventive measures, to improve patient safety, to minimize treatment cost and to design appropriate treatment strategy for pressure ulcer patients with spinal cord injury. In addition, the finding serves as alarming to health care professional to give a focus on the application of standardized care and represents a marker of quality of care.

Although this meta-analysis conducted with the use of comprehensive search strategy to incorporate the studies across a globe and all the included studies were observational study, there are some limitations that need to be considered in the future research: First, only English articles were considered; Second, this study do not identify the predictors of pressure ulcers among patients with spinal cord injury; Third, all included studies were reported hospital-based data.

## Conclusion and recommendations

This systematic review and meta-analysis revealed that about one in three patients with spinal cord injury had pressure ulcers. This implies that the overall global magnitude of pressure ulcer is relatively high. Therefore, policymakers (FMoH) and other concerned bodies need to give special attention to improve health care delivery for patient with spinal cord injury and reduce the risk of ulceration. Situation based interventions and country context specific effective preventive strategies should be developed to reduce the burden of pressure ulcers among patients with spinal cord injury and to improve the overall quality of healthcare service at large. Furthermore, further research is needed to identify associated factors for the development of pressure ulcers among patient with spinal cord injury.

## Data Availability

The data analyzed during the current meta-analysis is available from the corresponding author on reasonable request.

## Abbreviations

CI: Confidence Interval
FMoH: Federal Minster of Health
NPUAP: National Pressure Ulcer Advisory Panel
PU: Pressure Ulcer
PRISMA: Preferred Reporting Items for Systematic Reviews and Meta-Analyses
SCI: Spinal Cord Injury

## Declaration

### Ethics approval and consent to participate

Not applicable.

### Consent for publication

Not applicable.

### Authors’ contributions

WS and TY developed the protocol and involved in the design, selection of study, data extraction, and statistical analysis and developing the initial drafts of the manuscript. YA,HM and TY involved in data extraction, quality assessment, statistical analysis and revising. WS and YA prepared the final draft of the manuscript. All authors read and approved the final draft of the manuscript.

## Acknowledgements

We would like to thank all authors of studies included in this meta-analysis.

## Reference

1. Jang Y, Wang Y-H, Wang J-D: Return to work after spinal cord injury in Taiwan: the contribution of functional independence. Archives of physical medicine and rehabilitation 2005, 86(4):681–686.

2. Dumont RJ, Okonkwo DO, Verma S, Hurlbert RJ, Boulos PT, Ellegala DB, Dumont AS: Acute spinal cord injury, part I: pathophysiologic mechanisms. Clinical neuropharmacology 2001, 24(5):254–264.

3. White BA, Dea N, Street JT, Cheng CL, Rivers CS, Attabib N, Kwon BK, Fisher CG, Dvorak MF: The economic burden of urinary tract infection and pressure ulceration in acute traumatic spinal cord injury admissions: evidence for comparative economics and decision analytics from a matched case-control study. Journal of neurotrauma 2017, 34(20):2892–2900.

4. Rahimi-Movaghar V, Moradi-Lakeh M, Rasouli MR, Vaccaro AR: Burden of spinal cord injury in Tehran, Iran. Spinal Cord 2010, 48(6):492.

5. Jazayeri SB, Beygi S, Shokraneh F, Hagen EM, Rahimi-Movaghar V: Incidence of traumatic spinal cord injury worldwide: a systematic review. European spine journal 2015, 24(5):905–918.

6. Li C, DiPiro N, Cao Y, Szlachcic Y, Krause J: The association between metabolic syndrome and pressure ulcers among individuals living with spinal cord injury. Spinal cord 2016, 54(11):967.

7. Iyun AO, Malomo AO, Oluwatosin OM, Ademola SA, Shokunbi MT: Pattern of presentation of pressure ulcers in traumatic spinal cord injured patients in University College Hospital, Ibadan. International wound journal 2012, 9(2):206–213.

8. Mathew A, Samuelkamaleshkumar S, Radhika S, Elango A: Engagement in occupational activities and pressure ulcer development in rehabilitated South Indian persons with spinal cord injury. Spinal cord 2013, 51(2):150.

9. Kruger EA, Pires M, Ngann Y, Sterling M, Rubayi S: Comprehensive management of pressure ulcers in spinal cord injury: current concepts and future trends. The journal of spinal cord medicine 2013, 36(6):572–585.

10. Black J, Baharestani MM, Cuddigan J, Dorner B, Edsberg L, Langemo D, Posthauer ME, Ratliff C, Taler G: National Pressure Ulcer Advisory Panel’s updated pressure ulcer staging system. Advances in skin & wound care 2007, 20(5):269–274.

11. Ghaisas S, Pyatak EA, Blanche E, Blanchard J, Clark F: Lifestyle changes and pressure ulcer prevention in adults with spinal cord injury in the pressure ulcer prevention study lifestyle intervention. American Journal of Occupational Therapy 2015, 69(1):6901290020p6901290021–6901290020p6901290010.

12. Gorecki C, Brown JM, Nelson EA, Briggs M, Schoonhoven L, Dealey C, Defloor T, Nixon J, group EQoLPUP: Impact of pressure ulcers on quality of life in older patients: a systematic review. Journal of the American Geriatrics Society 2009, 57(7):1175–1183.

13. Lala D, Dumont FS, Leblond J, Houghton PE, Noreau L: Impact of pressure ulcers on individuals living with a spinal cord injury. Archives of physical medicine and rehabilitation 2014, 95(12):2312–2319.

14. Krause JS, Saunders LL: Health, secondary conditions, and life expectancy after spinal cord injury. Archives of physical medicine and rehabilitation 2011, 92(11):1770–1775.

15. Krause JS, Zhai Y, Saunders LL, Carter RE: Risk of mortality after spinal cord injury: an 8-year prospective study. Archives of physical medicine and rehabilitation 2009, 90(10):1708–1715.

16. Le Fort M, Espagnacq M, Albert T, Lefèvre C, Perrouin-Verbe B, Ravaud J-F: Risk of pressure ulcers in tetraplegic people: a French survey crossing regional experience with a long-term follow-up. European journal of public health 2018, 28(6):993–999.

17. Chan BC, Nanwa N, Mittmann N, Bryant D, Coyte PC, Houghton PE: The average cost of pressure ulcer management in a community dwelling spinal cord injury population. International wound journal 2013, 10(4):431–440.

18. Van der Wielen H, Post M, Lay V, Gläsche K, Scheel-Sailer A: Hospital-acquired pressure ulcers in spinal cord injured patients: time to occur, time until closure and risk factors. Spinal Cord 2016, 54(9):726.

19. Zakrasek E, Creasey G, Crew J: Pressure ulcers in people with spinal cord injury in developing nations. Spinal Cord 2015, 53(1):7.

20. Eslami V, Saadat S, Arejan RH, Vaccaro A, Ghodsi S, Rahimi-Movaghar V: Factors associated with the development of pressure ulcers after spinal cord injury. Spinal Cord 2012, 50(12):899.

21. Chen Y, DeVivo MJ, Jackson AB: Pressure ulcer prevalence in people with spinal cord injury: age-period-duration effects. Archives of physical medicine and rehabilitation 2005, 86(6):1208–1213.

22. Idowu O, Yinusa W, Gbadegesin S, Adebule G: Risk factors for pressure ulceration in a resource constrained spinal injury service. Spinal cord 2011, 49(5):643.

23. Verschueren J, Post M, De Groot S, Van der Woude L, Van Asbeck F, Rol M: Occurrence and predictors of pressure ulcers during primary in-patient spinal cord injury rehabilitation. Spinal cord 2011, 49(1):106.

24. Joseph C, Wikmar LN: Prevalence of secondary medical complications and risk factors for pressure ulcers after traumatic spinal cord injury during acute care in South Africa. Spinal Cord 2016, 54(7):535.

25. Correa G, Fuentes M, Gonzalez X, Cumsille F, Pineros J, Finkelstein J: Predictive factors for pressure ulcers in the ambulatory stage of spinal cord injury patients. Spinal cord 2006, 44(12):734.

26. Krause JS, Vines CL, Farley TL, Sniezek J, Coker J: An exploratory study of pressure ulcers after spinal cord injury: relationship to protective behaviors and risk factors. Archives of physical medicine and rehabilitation 2001, 82(1):107–113.

27. Gould LJ, Olney CM, Nichols JS, Block AR, Simon RM, Guihan M: Spinal cord injury survey to determine pressure ulcer vulnerability in the outpatient population. Medical hypotheses 2014, 83(5):552–558.

28. Fazel FS, Derakhshanrad N, Yekaninejad MS, Vosoughi F, Derakhshanrad A, Saberi H: Predictive value of braden risk factors in pressure ulcers of outpatients with spinal cord injury. Acta Medica Iranica 2018:56–61.

29. Pancorbo-Hidalgo PL, et al.: Risk assessment scales for pressure ulcer prevention: a systematic review. Journal of advanced nursing 2006, 54(1):p. 94–110.

30. Shea BJ, Reeves BC, Wells G, Thuku M, Hamel C, Moran J, Moher D, Tugwell P, Welch V, Kristjansson E: AMSTAR 2: a critical appraisal tool for systematic reviews that include randomised or non-randomised studies of healthcare interventions, or both. Bmj 2017, 358:j4008.

31. Borenstein M, Hedges L, Higgins J, Rothstein H: Introduction to Meta-Analysis John Wiley & Sons. Ltd, Chichester, UK 2009.

32. Egger M, Davey-Smith G, Altman D: Systematic reviews in health care: meta-analysis in context: John Wiley & Sons; 2008.

33. Rücker G, Schwarzer G, Carpenter J: Arcsine test for publication bias in meta-analyses with binary outcomes. Statistics in medicine 2008, 27(5):746–763.

34. StataCorp L: Stata statistical software (version release 14). College Station, TX: Author 2015.

35. Liberati A, Altman DG, Tetzlaff J, Mulrow C, Gøtzsche PC, Ioannidis JP, Clarke M, Devereaux PJ, Kleijnen J, Moher D: The PRISMA statement for reporting systematic reviews and meta-analyses of studies that evaluate health care interventions: explanation and elaboration. PLoS medicine 2009, 6(7):e1000100.

36. Brienza D, Krishnan S, Karg P, Sowa G, Allegretti A: Predictors of pressure ulcer incidence following traumatic spinal cord injury: a secondary analysis of a prospective longitudinal study. Spinal cord 2018, 56(1):28.

37. DeJong G, Hsieh C-HJ, Brown P, Smout RJ, Horn SD, Ballard P, Bouchard T: Factors associated with pressure ulcer risk in spinal cord injury rehabilitation. American journal of physical medicine & rehabilitation 2014, 93(11):971–986.

38. Garber SL, Rintala DH, Hart KA, Fuhrer MJ: Pressure ulcer risk in spinal cord injury: predictors of ulcer status over 3 years. Archives of physical medicine and rehabilitation 2000, 81(4):465–471.

39. Krishnan S, Karg PE, Boninger ML, Brienza DM: Association between presence of pneumonia and pressure ulcer formation following traumatic spinal cord injury. The journal of spinal cord medicine 2017, 40(4):415–422.

40. Richard-Denis A, Thompson C, Bourassa-Moreau É, Parent S, Mac-Thiong J-M: Does the Acute Care Spinal Cord Injury Setting Predict the Occurrence of Pressure Ulcers at Arrival to Intensive Rehabilitation Centers? American Journal of Physical Medicine & Rehabilitation 2016, 95(4):300–308.

41. Saunders LL, Krause JS, Acuna J: Association of race, socioeconomic status, and health care access with pressure ulcers after spinal cord injury. Archives of physical medicine and rehabilitation 2012, 93(6):972–977.

42. Ash D: An exploration of the occurrence of pressure ulcers in a British spinal injuries unit. Journal of Clinical Nursing 2002, 11(4):470–478.

43. Haisma JA, Van Der Woude LH, Stam HJ, Bergen MP, Sluis TA, Post MW, Bussmann JB: Complications following spinal cord injury: occurrence and risk factors in a longitudinal study during and after inpatient rehabilitation. Journal of rehabilitation medicine 2007, 39(5):393–398.

44. Raghavan P, Raza W, Ahmed Y, Chamberlain M: Prevalence of pressure sores in a community sample of spinal injury patients. Clinical rehabilitation 2003, 17(8):879–884.

45. Scheel-Sailer A, Wyss A, Boldt C, Post M, Lay V: Prevalence, location, grade of pressure ulcers and association with specific patient characteristics in adult spinal cord injury patients during the hospital stay: a prospective cohort study. Spinal cord 2013, 51(11):828.

46. Sheerin F, Gillick A, Doyle B: Pressure ulcers and spinal-cord injury: incidence among admissions to the Irish national specialist unit. Journal of wound care 2005, 14(3):112–115.

47. Kovindha A, Kammuang-Lue P, Prakongsai P, Wongphan T: Prevalence of pressure ulcers in Thai wheelchair users with chronic spinal cord injuries. Spinal cord 2015, 53(10):767.

48. Tchvaloon E, Front L, Gelernter I, Ronen J, Bluvshtein V, Catz A: Survival, neurological recovery and morbidity after spinal cord injuries following road accidents in Israel. Spinal cord 2008, 46(2):145.

49. Taghipoor KD, Arejan RH, Rasouli MR, Saadat S, Moghadam M, Vaccaro AR, Rahimi-Movaghar V: Factors associated with pressure ulcers in patients with complete or sensory-only preserved spinal cord injury: is there any difference between traumatic and nontraumatic causes? Journal of Neurosurgery: Spine 2009, 11(4):438–444.

50. Löfvenmark I, Wikmar LN, Hasselberg M, Norrbrink C, Hultling C: Outcomes 2 years after traumatic spinal cord injury in Botswana: a follow-up study. Spinal cord 2017, 55(3):285.

51. Teena Chopra MD M, Dror Marchaim MD, Reda A. Awali MD, MPH,,, Miriam Levine MD SSM, Indu K. Chalana MD, Farah Ahmed MD Risk factors and acute in-hospital costs for infected pressure ulcers among gunshot-spinal cord injury victims in southea stern Michigan. american Journal of infection control 2016, 44(3):315–319.

52. Fazel FS DN, Yekaninejad MS, Vosoughi F, Derakhshanrad A, Saberi H. : Predictive value of braden risk factors in pressure ulcers of outpatients with spinal cord injury. Acta Medica Iranica 2018, 56(1):56–61.

53. Al Mutairi KB, & Hendrie, D. : Global incidence and prevalence of pressure injuries in public hospitals:A systematic review. Wound Medicine, 2018, 22:23–31.

54. Xakellis Jr GC, Frantz RA, Lewis A, Harvey P: Translating pressure ulcer guidelines into practice: it’s harder than it sounds. Advances in skin & wound care 2001, 14(5):249–258.

